# The expansion and severity of chronic MS lesions follows a periventricular gradient

**DOI:** 10.1101/2021.12.20.21268136

**Authors:** Samuel Klistorner, Michael H Barnett, Stuart L Graham, Chenyu Wang, Alexander Klistorner

## Abstract

**Background and Objectives:** Expansion of chronic lesions in MS patients and recently described CSF-related gradient of tissue damage are linked to microglial activation. The aim of the current study was to investigate whether lesion expansion is associated with proximity to ventricular CSF spaces.

**Methods:** Pre- and post-gadolinium 3D-T1, 3D FLAIR and diffusion tensor images were acquired from 36 RRMS patients. Lesional activity was analysed between baseline and 48 months at different distances from the CSF using successive 1-mm thick concentric rings radiating from the ventricles.

**Results:** Voxel-based analysis of the rate of lesion expansion demonstrated a clear periventricular gradient decreasing away from the ventricles. This was particularly apparent when lesions of equal diameter were analysed. Periventricular lesional tissue showed higher degree of tissue distraction at baseline that significantly increased during follow-up in rings close to CSF. This longitudinal change was proportional to degree of lesion expansion. Lesion-wise analysis revealed a gradual, centrifugal decrease in the proportion of expanding lesions from the immediate periventricular zone.

**Discussion:** Our data suggest that chronic white matter lesions in close proximity to the ventricles are more destructive, show a higher degree of expansion at the lesion border and accelerated tissue loss in the lesion core.

## Introduction

Axonal loss is now accepted as the major cause of irreversible neurological disability in MS. While acute inflammatory demyelination, typically manifested as new white matter lesions, is thought to be a principal cause of axonal transection and subsequent axonal degeneration, slow-burning inflammatory demyelination at the edge of chronic lesions and its MRI equivalent (the expansion of chronic MS lesions) has been suggested as another major factor contributing to disease evolution, including progressive neurodegeneration, brain atrophy and disability worsening.^1,2,3,4^ Although the mechanisms responsible for this phenomenon are still poorly understood, microglial activation and myelin breakdown play a central role.^5,6^

A CSF-related gradient of tissue injury has recently been described in both normal-appearing white matter (NAWM) and chronic lesions in patients with MS.^7,8,9,10,11^ White matter magnetization transfer ratio (MTR) and Mean Diffusivity (MD) abnormalities were reported around the ventricles, with the most severe abnormalities bordering the ventricular surface. Akin to pathomechanisms thought to be involved in slow lesion expansion, the periventricular injury gradient is linked to innate immune cell activation^12^ and seems to be independent of formation of new lesions.^3,7^

Therefore, the aim of the current study was to investigate whether slow-burning inflammation at the rim of chronic MS lesions (assessed by the degree of lesion expansion) is associated with proximity to the CSF; and to explore a potential relationship between lesion topography and the degree of progressive tissue injury within the lesional core (as measured by T1 hypointensity and MD).

## Methods

The study was approved by University of Sydney and Macquarie University Human Research Ethics Committees and followed the tenets of the Declaration of Helsinki. Written informed consent was obtained from all participants.

### Subjects

Fifty consecutive patients with established RRMS, defined according to the revised McDonald 2010 criteria^13^, were enrolled in a study. Patients underwent MRI scans and clinical assessment at 0 months, 12 months and 60 months. The main analysis was performed between 12 months (termed “baseline”) and 60 months (termed “follow-up”), while scans performed at 0 months (termed “pre-study scans”) were used to identify (and exclude) newly developed lesions at the start of the study.^3^

### MRI protocol

MRI was performed using a 3T GE Discovery MR750 scanner (GE Medical Systems, Milwaukee, WI). Specific parameters and MRI image processing are presented in Supplementary material.

### Generation of periventricular

Ventricular CSF was semi-automatically segmented using JIM 7 software (Xinapse Systems, Essex, UK) on baseline scans by a trained analyst. The white matter of each subject was then segmented into 1-mm-thick concentric rings starting at the ventricular margin, as described in^11^ (Fig. 1). The first two rings were excluded from analysis to limit partial volume effects from CSF, as previously suggested.^8,10^

**Fig. 1.**
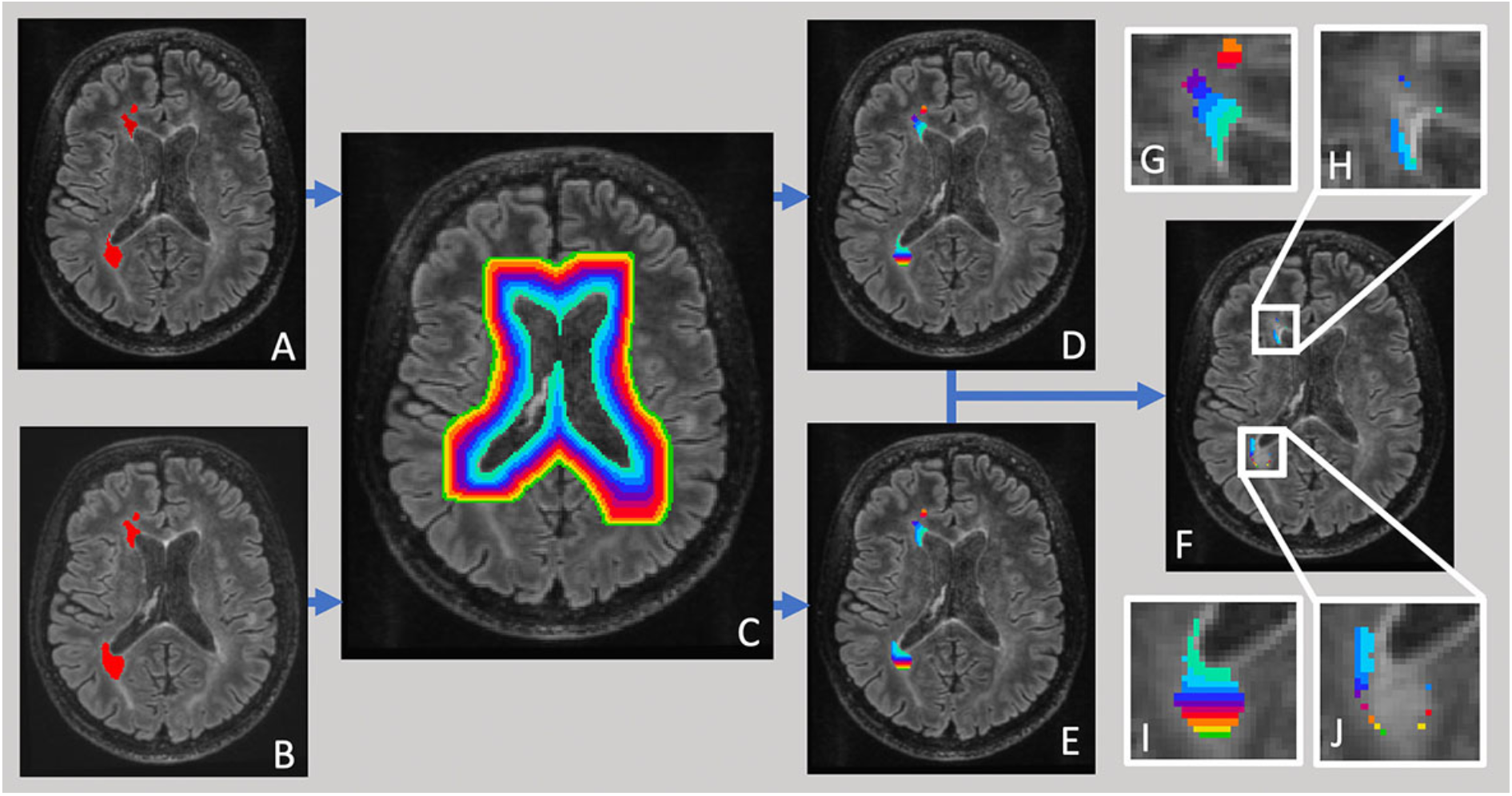
Pipeline of ring-based lesion analysis. Lesions at baseline (a) and follow-up (b) were semi-automatically segmented and intersected with concentric periventricular rings of 1 mm thickness (c, d, e). Average lesion expansion was calculated for each ring as difference between follow-up and baseline lesion masks (f). Inserts magnify individual lesions (g, i-lesion mask at baseline intersected with ring pattern), demonstrating larger expansion in rings close to ventricles (h, j-lesion expansion).

All voxels within individual periventricular rings were averaged together and a single MD or T1 hypointensity value was used to estimate tissue damage within lesional or NAWM volume.

For ring-based analysis, the number of voxels at baseline and follow-up within a single ring were estimated. The rate of lesion expansion within the ring was then calculated as a relative change in number of lesional voxels occurring during the follow-up.^3^ To reduce the disproportionally large impact of small lesions on calculation of CSF-related gradient of lesion expansion, the weight corresponding to number of baseline lesional voxels in each ring was assigned to each lesion, as recently suggested.^12,10^

For lesion-based analysis, the “center of mass” for each lesion was calculated using the weighted average of lesional voxels distributed across all the rings. Lesions with a “center of mass” within the first 10 rings were used for analysis.

### Statistics

Statistical analysis was performed using SPSS 22.0 (SPSS, Chicago, IL, USA). Pearson correlation coefficient was used to measure statistical dependence between two numerical variables. P < 0.05 was considered statistically significant. Comparisons between groups were made using Student *t*-test. Longitudinal changes were assessed using paired two-sample *t*-test. Fisher’s exact test was used for categorical data.

## Results

Thirty-six patients completed the study. Demographic data is presented in Table 1.

**Table 1.** Demographic data (^1^ patients remain on the same DMT for the bulk of the study). Two patients received no DMT through the course of the study.

|  | Demographic Variables |
| --- | --- |
| Total number of patients | 36 |
| Age at enrolment, years | 43.3±10.2 |
| Gender | 14M/22F |
| Disease duration at enrolment, years | 5.5±2.8 |
| EDSS at baseline (median) | 1.0+1.1 |
| No. of patients on stable treatment<sup>1</sup> | 34 |
| Betaferon | 2 |
| Avonex | 5 |
| Rebif | 1 |
| Glatiramer acetate | 5 |
| Fingolimod | 10 |
| Dimethyl fumarate | 3 |
| Natalizumab | 8 |

Lesion volume at baseline exponentially decayed from the periventricular region outwards (Suppl Fig. 1a). Therefore, only data from the first ten rings was analysed. There were 349 lesions fully or partially within the first 10 rings.

### Periventricular gradient of lesion expansion and severity

The averaged (ring-wise) volume of expanding chronic lesions and new (both free-standing and confluent) lesions is plotted in eFig.1. Lesions starting at any ring and occupying any number of rings are included. New lesional activity demonstrated a relatively small uniformly distributed volume, while expanding volume of chronic lesions sharply increases toward the CSF.

The combined analysis of lesion expansion rate, grouped by the distance from CSF, is presented in Fig. 2a (blue dots). The rate of expansion shows linear reduction with increasing distance from the ventricles.

**Fig. 2.**
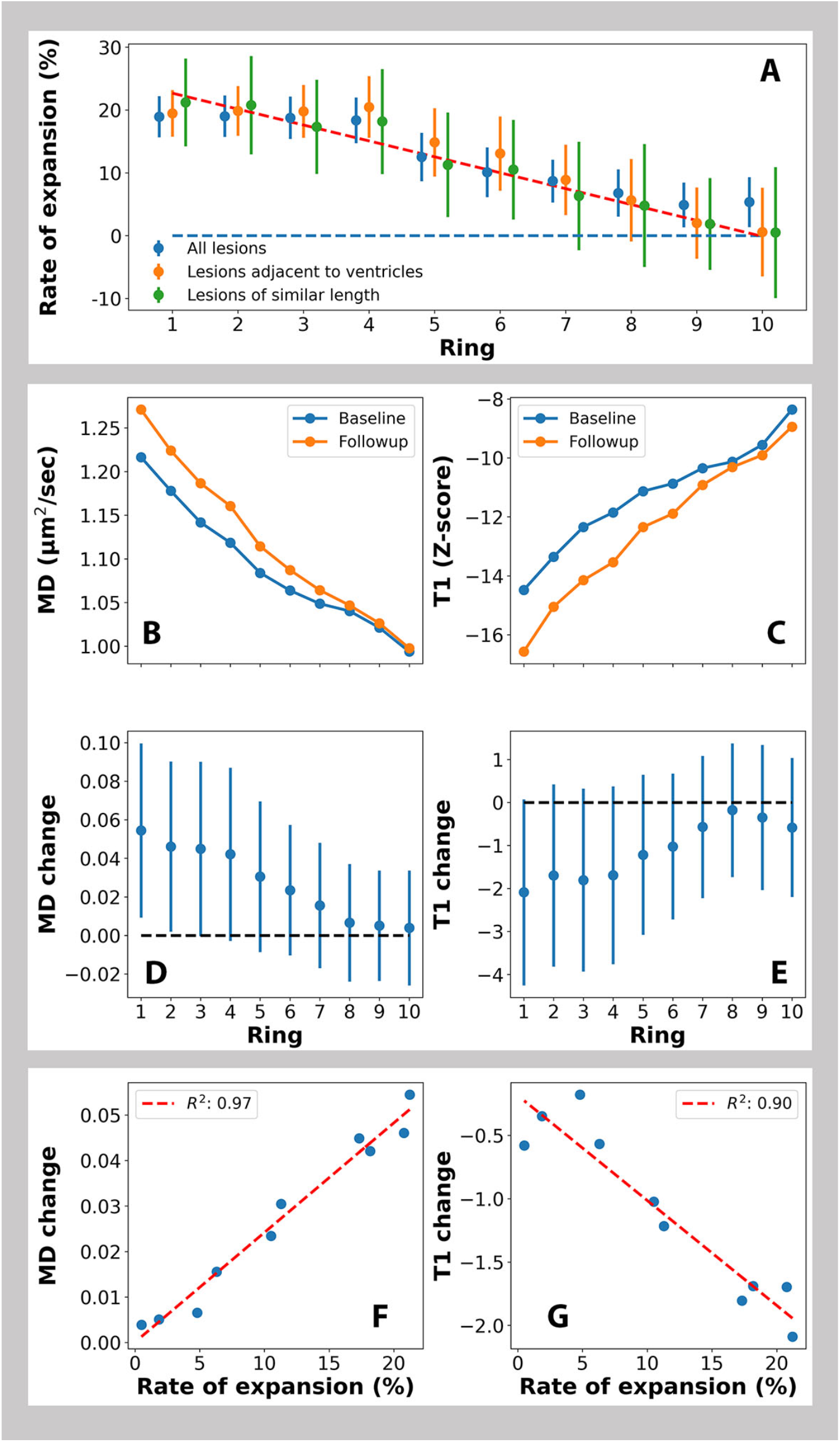
Voxel-based analysis of lesion expansion and severity at increasing distance from the ventricles. a: CSF-related gradient of lesion expansion, Error bars represent 2 SE. Blue dots: all lesions. Orange dots: lesions adjacent to the ventricles Green dots: lesions adjacent to the ventricles and of similar length (red line represents best linear fit). b-e: Ring-wise analysis of MD and T1 intensity within lesions: b. Baseline and follow-up MD values c. MD change during study period d. Baseline and follow-up T1 intensity values e. T1 intensity change during study period Error bars represent 2 SE. f-g: Correlation between the rate of lesional volume expansion at different periventricular rings and corresponding values of: f. Mean Diffusivity (MD units are μm^2^/ms) g. T1 hypointensity (T1 units are z-score).

In accordance with a previous study^14^, we found that white matter lesions are more prevalent adjacent to the ventricles, constituting the bulk of lesional volume (91%) within first 10 rings. Therefore, as expected, when analysis was restricted to those lesions (144 in total) the pattern of lesion expansion did not materially change (Fig. 2a, orange dots)

The size of lesions adjacent to the ventricles exhibited significant heterogeneity, occupying only the first two or three rings in some lesions, or extending 20 mm or beyond from the ventricle border in others (Suppl. Fig. 2). Therefore, we further limited the analysis to lesions covering an approximately equal number of rings (10-13 rings) in order to minimise the effect of variable lesion size. There were 26 lesions from 16 subjects conforming to this size. Longitudinal analysis of these lesions (Fig. 2a, green dots) demonstrated even more linear CSF-related gradient of ring-wise expansion (correlation between average rate of expansion and distance from the CSF, r = -0.97, p<0.0001).

### Progressive change in lesional T1 hypointensity and MD tracks the periventricular lesion expansion gradient

This analysis was performed using 26 lesions of similar length, as described above. Periventricular lesional tissue demonstrated a higher MD and lower T1 intensity value at baseline indicating a greater degree of tissue damage within lesions closer to the CSF (Fig. 2b, c). In addition, longitudinal analysis of lesional tissue also revealed a significant increase of MD and reduction of T1 intensity in rings close to CSF, suggesting a periventricular gradient of progressive axonal loss inside lesions (Fig. 2d, e). This change was highly proportional to the degree of lesion expansion (r=0.97, p<0.001 and r=0.89, p<0.001 for MD and T1 respectively, Fig. 2f, g). T1 intensity and MD metrics at baseline and follow-up and significance of change in these values in individual rings are presented in Table. 2

**Table 2.** Baseline and follow-up values of lesion volume, MD and T1 intensity in individual periventricular rings. Significant differences are shown in bold. MD units are μm^2^/ms, T1 units are z-score.

|  | Baseline volume | Follow-up volume | Vol. P value | Baseline MD | Follow-up MD | MD P value | Baseline T1 | Follow-up T1 | T1 P value |
| --- | --- | --- | --- | --- | --- | --- | --- | --- | --- |
| Ring 1 | 8975 | 10879 | **0.018** | 1.16 | 1.2 | **0.007** | -13.06 | -14.5 | **0.022** |
| Ring 2 | 7500 | 9058 | **0.014** | 1.14 | 1.18 | **0.012** | -12.62 | -13.93 | **0.049** |
| Ring 3 | 5913 | 6937 | **0.013** | 1.11 | 1.15 | **0.008** | -11.51 | -13.13 | **0.024** |
| Ring 4 | 4227 | 4995 | **0.012** | 1.08 | 1.12 | **0.006** | -10.89 | -12.46 | **0.023** |
| Ring 5 | 3262 | 3630 | **0.018** | 1.05 | 1.08 | **0.01** | -10.6 | -11.94 | **0.021** |
| Ring 6 | 2331 | 2576 | **0.043** | 1.04 | 1.06 | 0.084 | -10.44 | -11.44 | 0.071 |
| Ring 7 | 1853 | 1970 | 0.181 | 1.04 | 1.06 | 0.174 | -10.43 | -11.14 | 0.209 |
| Ring 8 | 1457 | 1527 | 0.331 | 1.03 | 1.04 | 0.439 | -10.35 | -10.61 | 0.596 |
| Ring 9 | 1129 | 1150 | 0.581 | 1.01 | 1.02 | 0.369 | -9.35 | -9.88 | 0.404 |
| Ring 10 | 800 | 804 | 0.92 | 0.97 | 0.98 | 0.586 | -8.02 | -8.53 | 0.502 |

### Lesion-wise analysis of the periventricular gradient of lesion expansion and severity

To examine the periventricular gradient between individual lesions, a “center of mass” for each lesion was calculated and assigned to periventricular rings 1 mm thick extending up to 10 mm from CSF, as described in Methods. There were 280 lesions in total, 113 (40%) of which were expanding and 167 of which were stable or shrinking.

Lesions with a “center of mass” within the first 2 rings from the lateral ventricles were grouped together into a periventricular WM cluster, lesions with the “center of mass” in rings 3-4 were grouped into intermediate WM cluster and lesions with the “center of mass” between 5^th^ and 10^th^ rings were grouped into deep WM cluster ^8^. The proportion of expanding lesions for each cluster was computed. The result of this analysis, presented in Fig. 3, demonstrated a gradual decrease in the proportion of expanding lesions from periventricular area towards the deep NAWM. The proportion of expanding lesions within the periventricular area was significantly higher than within the deep white matter group (Chi-Square Fisher’s exact test=0.031).

**Fig. 3.**
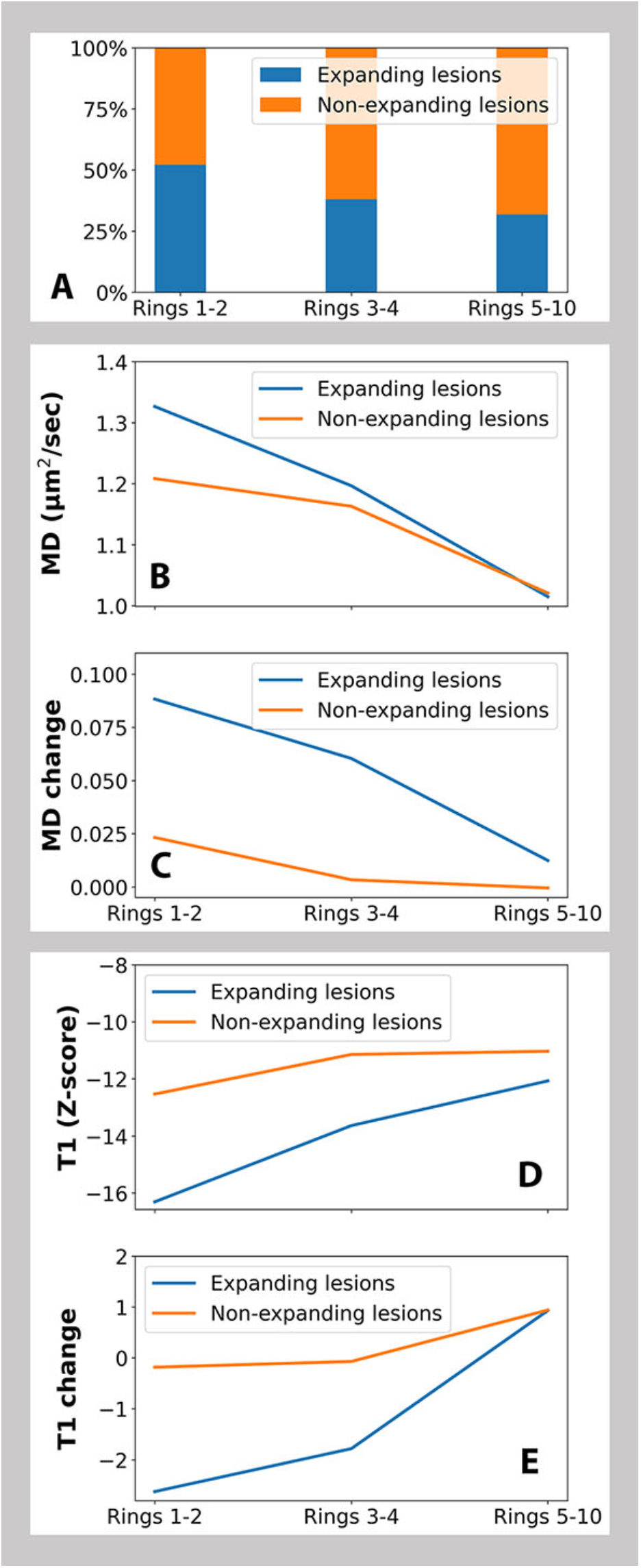
Lesion-based analysis a. Proportion of expanding lesions in periventricular, intermediate, and deep WM clusters. b. Baseline MD values in expanding and non-expanding lesions in periventricular, intermediate, and deep WM lesion clusters. c. Change of MD in expanding and non-expanding lesions in periventricular, intermediate, and deep WM clusters during follow-up d. Baseline values of T1 hypointensity in expanding and non-expanding lesions periventricular, intermediate, and deep WM clusters. e. Change of T1 hypointensity in expanding and non-expanding lesions in periventricular, intermediate, and deep WM clusters during follow-up

The baseline CSF-related gradient of MD and T1 hypointensity was clearly more apparent in expanding compared to non-expanding lesions. The value of MD and T1 hypointensity at baseline was significantly different between expanding and non-expanding lesions in the periventricular WM cluster, but not in the intermediate or deep WM clusters (Fig. 3 b and d, Table. 3). Moreover, the CSF-related gradient of progressive change of MD and T1 hypointensity during follow-up was observed only in expanding, but not in non-expanding lesions. The difference between the two groups was highly significant in periventricular and intermediate WM clusters, but not in the deep WM group (Fig. 3 c and e. Table. 3).

**Table 3.** Baseline values and progressive change of MD and T1 hypointensity in expanding vs non-expanding lesions in periventricular, intermediate and deep WM clusters. Significant differences are shown in bold. MD units are μm^2^/ms, T1 units are z-score.

|  | MD Expanding | MD Non-expanding | MD P value | T1 Expanding | T1 Non-expanding | T1 P value |
| --- | --- | --- | --- | --- | --- | --- |
| **Baseline** |  |  |  |  |  |  |
| Ring 1-2 | 1.27 | 1.2 | **0.013** | -15.99 | -12.86 | **0.006** |
| Ring 3-4 | 1.09 | 1.1 | 0.754 | -11.6 | -11.79 | 0.872 |
| Ring 5-10 | 0.99 | 1 | 0.704 | -11.03 | -10.27 | 0.454 |
| **Progressive change** |  |  |  |  |  |  |
| Ring 1-2 | 0.08 | 0.02 | **>0.001** | -2.02 | -0.6 | **0.01** |
| Ring 3-4 | 0.07 | 0.01 | **>0.001** | -1.58 | 0.39 | **0.004** |
| Ring 5-10 | 0.01 | 0 | 0.329 | 0.25 | 0.45 | 0.726 |

### Periventricular gradient of abnormality in NAWM at different distances from the lesion border

The strong periventricular MD gradient observed within chronic MS lesions raises the possibility that the previously reported CSF-related abnormality in NAWM may be driven by lesional changes (for instance, by Wallerian degeneration of the fibers transected at the lesion rim as a result of slow-burning inflammation^15^). Therefore, to investigate whether the MD gradient in NAWM declines with increasing distance from the lesion border, we examined diffusivity change in five consecutive perilesional rings (each 2 voxel thick) expanding into NAWM.

The results are presented in Fig. 4a, demonstrating that, apart from the first ring (i.e. closest 2 voxels to the lesion border), no significant difference in the NAWM MD gradient was observed. Even the removal of two patients with significant new lesional activity did not materially change the result.

**Fig. 4.**
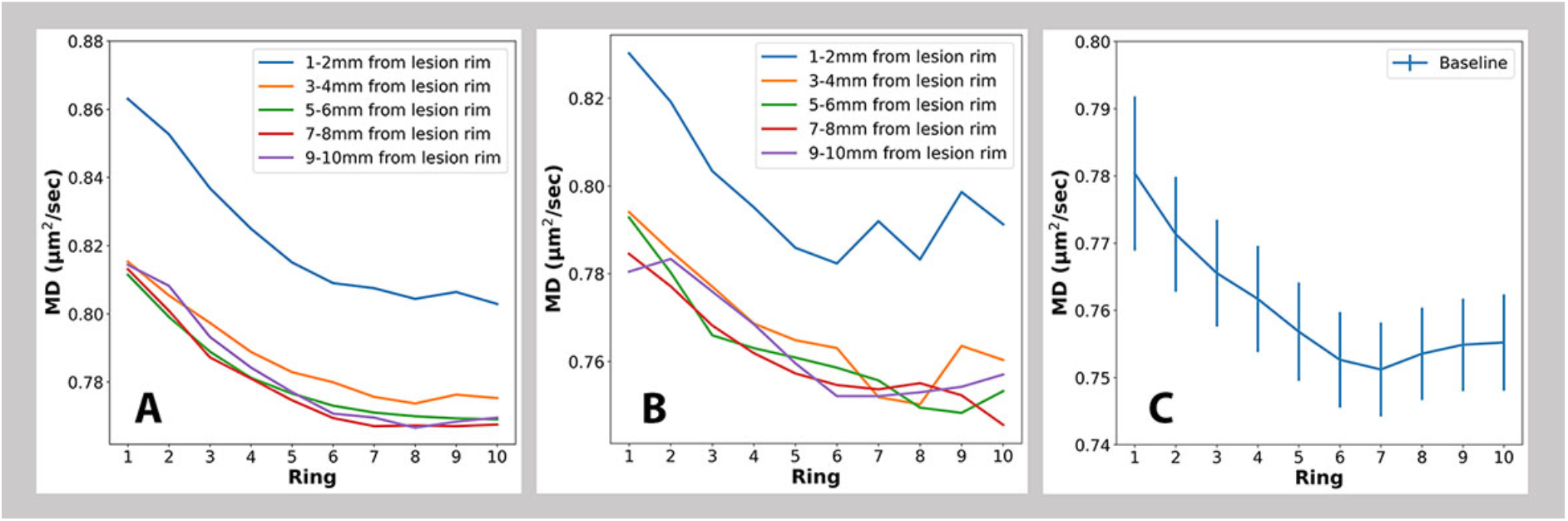
NAWM analysis. a. Ring-wise profile of MD in NAWM at various distances from lesion border including all patients b. Ring-wise profile of MD in NAWM at various distance from CSF obtained from 9 patients with minimal periventricular lesion load. c.Ring-wise MD profile in deep NAWM including all patients.

A similar gradient was also observed when data from 9 patients with minimal periventricular lesion load was analysed separately (Suppl Fig 4b).

Moreover, in a separate analysis, the lesion mask was expanded by 10 voxels in order to exclude the lesion surroundings of up to 1 cm. MD analysis of the remaining NAWM voxels, grouped into periventricular rings, showed a clear declining gradient of microstructural damage severity extending from the ventricular border (Fig. 4c).

## Discussion

### Periventricular gradient of expansion and severity in chronic MS lesions

We explored the relationship between the rate of expansion in chronic MS lesions and distance from the adjacent CSF spaces. We found that the rate of lesion expansion in patients with RRMS is inversely proportional to the distance from the ventricular system. This CSF-based gradient was observed both within lesions, as well as between individual lesions. Examining periventricular lesions of similar size revealed a high degree of correlation between reduction of the rate of lesion expansion and increasing distance from CSF, while grouped analysis of individual lesions demonstrated a significantly higher proportion of expanding lesions in close proximity to the ventricles compared to the deep white matter. While earlier work has indicated that the anatomical distribution of expanding lesions follows the common periventricular pattern of lesion distribution in MS,^2,16^ this study is the first to demonstrate a direct relationship between the degree of chronic lesion expansion and the distance from the ventricular border, highlighting CSF-related dependency of the chronic inflammatory process. Providing that lesion expansion is an imaging equivalent of slow-burning inflammation at the lesion rim, our results suggest that inflammatory activity at the rim of chronic lesions is also dependent on the distance from CSF.

Moreover, baseline voxel-based lesion analysis segmented by periventricular rings demonstrated a well-defined CSF-related gradient of MD and T1 hypointensity (i.e. significantly more abnormal values in periventricular lesional voxels compared to the area of the lesion more remote from CSF). Over the study period the rate of microstructural tissue damage within lesions, as measured by increase in MD and decrease in T1 intensity, followed a similar pattern, exhibiting the greatest change in close proximity to the CSF. Notably, the magnitude of the observed increase in MD and T1 hypointensity in the periventricular ring closest to the ventricles was highly comparable (4.0% and 3.8% respectively). However, the relatively small change in lesional MD and T1 observed values during the study is insufficient to account for the baseline gradient of both measures, which would require more than 15 years to develop based on the aforementioned rate of progression. Conversely, the average disease duration in our patient coghort was only 5.5 years. Taken together, these findings suggests that components of the lesion closer to the CSF are likely to incur more severe damage during acute inflammation associated with the lesion formation; and to accumulate greater damage during the chronic stage.

Lesion-based analysis corroborated the presence of CSF-related gradient of lesion expansion; and additionally revealed that the observed MD and T1 gradients of tissue injury, both at baseline and during the follow-up period, were primarily driven by the expanding lesions. The latter finding is in agreement with earlier studies showing the more destructive nature of chronic active lesions^17,18^

We recently demonstrated a close association between expansion of the lesion rim and progressive axonal damage inside the lesion and attributed this to Wallerian and retrograde degeneration of axons transected at the lesion rim.^3^ Similarly, progressive tissue damage inside slowly expanding lesions has been reported by others groups.^2,19^ The current study provides strong support for this notion by demonstrating that gradients of isotropic water diffusion and T1-intensity alteration inside lesions during the follow-up period are highly proportional to the periventricular gradient of lesion expansion, and, as a result, are likely to be secondary to slow-burning inflammation at the lesion rim.

### Periventricular gradient in NAWM

Our study confirmed the presence of a gradient of damage severity in NAWM.^11^ Moreover, the magnitude of MD increase in periventricular area of NAWM (∼5%, see Fig.7) is similar to the previously reported CSF-related MTR gradient.^7^ The lesion-independent nature of this gradient was further verified by the analysis of CSF-related MD curves at different distances from the lesion border, which (with exclusion of first 2 mm ring), showed very similar values. In addition, analysis of NAWM removed from the lesion border by more than 10 mm also yielded a discernible periventricular predominance of microstructural tissue damage. This is further supported by our demonstration of a CSF-related MD gradient in NAWM in a group of patients with no or very limited periventricular lesions, corroborating previous observations that periventricular gradients of the severity of tissue injury do not differ significantly between patients with or without white matter lesions.^10^

### Lesion expansion and NAWM periventricular gradient

The observed decline in the rate of lesion expansion and progressive microstructural damage within lesions as distance from the CSF increases, reported here; and gradient of lesion and NAWM tissue damage severity and microglial activation described in previous publications,^11,10,8^ have a number of common features that raise the possibility of shared pathophysiology.

First, the expansion of chronic lesions mirrors the CSF-associated gradient of microstructural abnormality in NAWM and lesional tissue^11,10,8,20,21^ and is most prominent in close proximity to the ventricles, declining markedly with increasing centrifugal distance. Furthermore, the baseline MD and T1 hypointensity values of lesional voxels, observed in our study, are more abnormal at the ventricular margin and recover with increasing distance from it. More importantly, a similar CSF-related gradient was also found when longitudinal assessment of lesional tissue injury was performed; namely, values of MD and T1 hypointensity significantly increased in rings close to CSF, indicating measurable progressive damage in periventricular lesional tissue during the follow-up period.

Second, both the rate of lesion expansion and periventricular gradient of tissue damage severity are associated with activation of microglia. Thus, a recent study has shown a periventricular gradient of microglial activation both in NAWM and MS lesions^12^; and post-mortem evidence has suggested the involvement of activated microglia in CSF-related tissue injury.^22^ Similarly, microglial activation has been implicated as a principal driver of slow-burning inflammation at the rim of chronic active lesions, which represents the pathophysiological equivalent of lesion expansion.^5,23,24,25^

Third, both the periventricular gradient of damage severity in NAWM and the rate of lesion expansion appear to be independent of new lesion formation, implying a pathological process distinct from acute inflammation. Multiple investigations have reported that the periventricular NAWM MTR gradient, which is of similar magnitude in patients with and without T2 lesions, is not directly related to lesion formation; may impact clinical outcomes independently from focal white matter lesions; and may, therefore, represent a novel therapeutic target.^7,10^ Similarly, the acute and chronic white matter lesions contribute independently to disease progression, implying that different mechanisms are likely to be responsible for acute and smouldering inflammation.^1,2,3,26,27^

Finally, the gradient of tissue severity damage in NAWM and the presence of chronic active lesions are more prominent in patients with progressive forms of MS.^11,14^

Therefore, it is plausible that some of the mechanisms proposed to explain the periventricular predominance of abnormalities in the NAWM, such as a neurotoxic effect of soluble CSF-derived factors,^12,28^ hypoxia^29^, reduced remyelinating capacity^30^ or increased compartmentalized inflammation^9^ in periventricular region and microglial activation^12^ are important mediators of the expansion of chronic lesions observed in our study.

Whatever the cause of the CSF-related tissue abnormality gradient in MS, its existence, corroborated by this study, helps to explain, at least to some extent, the heterogeneity of lesion expansion seen in MS patients.^3,19^ It appears that the closer the lesional voxels are to the CSF, the more likely they are to be affected by the factor(s) promoting slow-burning inflammation at the lesion rim and lesion expansion. The spatial extent of this effect, however, is limited to a few mm from the ventricular edge.

This study has some limitations, including a relatively small sample size and the absence of normal controls. A comparison with normal subjects, however, is only applicable to study of NAWM, which has been thoroughly undertaken in earlier investigations^7^. Another limitation is the absence of additional MRI techniques (eg MTR), which were not part of the acquisition protocol in this study. Based on recent publications, we believe that longitudinal studies combining PET, MTR, susceptibility and diffusivity imaging are needed to better understand role of innate immune cell activation in chronic inflammation associated with the lesion rim.^31^

## Conclusion

In conclusion, our data suggest that MS lesions in close proximity to the ventricles are not only more destructive, but also exhibit progressive expansion at the lesion rim and accelerated tissue degeneration in the lesion core. This CSF-related gradient appears to be present both within and between lesions, suggesting that the expansion of chronic lesions is not a lesion-driven process, but rather is associated with cytotoxic factors that are most active near the CSF and spread through the surrounding NAWM.

## Data Availability

All data produced in the present study are available upon reasonable request to the authors

## Supplementary material

### 1. Specific MRI parameters and image processing

The following MRI sequences were acquired:

a. Pre- and post-contrast (gadolinium) Sagittal 3D T1: GE BRAVO sequence, duration 4 min each, FOV 256mm, Slice thickness 1mm, TE 2.7ms, TR 7.2ms, Flip angle 12°, Pixel spacing 1mm. Acquisition Matrix (Freq x Phase) is 256×256, which results in 1mm isotropic acquisition voxel size. The reconstruction matrix is 256×256.
b. PFLAIR CUBE; GE CUBE T2 FLAIR sequence, duration 6 min, FOV 240mm, Slice thickness 1.2mm, Acquisition Matrix (Freq x Phase) 256×244, TE 163ms, TR 8000ms, Flip angle 90°, Pixel spacing 0.47 mm. The reconstruction matrix is 512×512.
c. PEcho-Planar Imaging based diffusion weighted MRI, duration 9 min (64-directions with2mm isotropic acquisition matrix, TR/TE = 8325/86 ms, b = 1000 s/mm^2^, number of b0s = 2).

### MRI image pre-processing

The baseline T1-weighted imaging was realigned to Anterior and Posterior Commissure (AC-PC) orientation. Using FLIRT (FSL, FMRIB Software Library), follow-up T1 images were co-registered to initial (month 0) AC-PC space by applying transformation matrices derived from linear co-registration between baseline AC-PC aligned brain and follow-up native T1 brain images. In parallel, diffusion MRI was corrected for motion and eddy-current distortion in FSL, then EPI susceptibility distortion was minimized by applying deformation maps generated from nonlinear co-registration between DWI b0 brain images and T1-weighted images at each time-point using ANTS (Advanced Normalization Tools). Subsequently, tensor reconstruction was performed in MRtrix3. Tensor and FLAIR images were then linearly co-registered to corresponding T1 AC-PC images at each timepoint.

### DTI data processing

Diffusion weighted MRI data (dMRI) were pre-processed using tools provided by the software suites MRtrix3 [1], FSL [2], and ANTs [3, 4]. Specifically, dMRI data were first denoised [5], then potential Gibbs-ringing artefacts were removed [6]. The dMRI data were then corrected for bias field inhomogeneities using the ANTs N4 algorithm [4]. a dMRI brain mask was then estimated using BET [7] and used as an input, alongside the dMRI data, to eddy [8] in order to correct for subject movement in the acquisition. Finally, phase distortion correction was applied using a non-linear registration method outlined below.

To correct for phase distortion within the dMRI data, first the brain was segmented from the corresponding T1w dataset, and a single B0 volume was extracted from the dMRI dataset. The T1w brain was then used as a mask to invert the contrast of the T1w image. A rigid-body registration was then performed on the inverted T1w image with the B0 volume as a target, which aligned the two images spatially. Non-linear registration was then performed using ANTs. The registration steps were comprised of a rigid body, then affine, and then SyN registration algorithm [3]. The transformations and warps calculated from the non-linear registration steps were then applied to the entire dMRI dataset to correct for phase distortion artefacts.

### Brain segmentation

Brain extraction and tissue segmentation was performed on T1 images using ANTS. Tissue segmentation was used to determine a white matter mask which was later used in conjunction with MD data. WhiteStripe normalisation tool (Cancer Imaging Phenomics Toolkit, CaPTk) was applied to T1 brain extracted images to normalise intensity between subjects ^17^.

#### Lesion identification and analysis

Individual lesions were identified on the co-registered T2 FLAIR images and semi-automatically segmented using JIM 7 software (Xinapse Systems, Essex, UK) on all scans by a trained analyst (Fig. 1b). Only lesions measuring larger than 50 mm^2^ were selected for analysis, as described previously ^3^.

Gadolinium enhancement within active MS lesions usually does not persist beyond 2 months, after which newly formed T2 hyperintense lesions continue to shrink in size for another 3-5 months, reflecting resolution of edema and, potentially, tissue repair including remyelination ^18^. Therefore, to minimize lesion changes related to evolution of newly developed lesions, both baseline gadolinium-enhancing lesions and newly formed baseline non-enhancing T2 lesions (i.e. lesions visible on baseline images, but not present on pre-study images) were excluded from the analysis.

Once selected, baseline and follow-up lesions were converted to binary mask and analysed by a fully automated in-house lesion progression algorithm written in Python, as previously reported ^3^. Briefly, the algorithm includes several steps, namely:

1. Lesion matching: Individual lesions were matched between baseline and follow-up time-points based on spatial connectedness (i.e. lesion overlap). All follow-up lesions that did not have a matched baseline counterpart were classified as “new free-standing lesions” and excluded from the analysis
2. Correction of follow-up lesions for brain atrophy: with relatively long periods of follow-up, brain atrophy frequently leads to a significant shift in the position of MS lesions^19^, which affects the accurate estimation of the lesion. Therefore, for each baseline-to-follow-up lesion pair the position of the follow-up lesion mask was adjusted for brain atrophy by implementing linear co-registration between the two masks using 6 degrees of freedom (rigid body) with limited rotation of around 5 degrees (FLIRT)^20^
3. Identification of new confluent lesions: performed by a custom-designed algorithm as described previously^3^
4. Calculation of chronic lesion volume change during follow-up: The final size of chronic lesions at follow-up was determined by subtracting confluent lesions from follow-up lesion mask. Subsequently, the difference between the remainder of the follow-up lesion mask and baseline lesion mask was used to calculate change in chronic lesion volume

Chronic lesions that demonstrated ≤10% volume change (positive or negative) were classified as stable (10% represents test–retest variability of manual lesion masking ^3^).

Lesions with >10% volume increase were classified as expanding, while lesions with < 10% volume decrease were classified as shrinking. Shrinking and stable lesions were grouped together into “non-expanding lesions”.

New free-standing lesions and new confluent lesions that developed during the follow-up were combined to compute the patient-wise volume of recent acute lesions.

The degree of tissue damage within chronic lesions was assessed by Mean Diffusivity (MD) ^19^ and T1 hypointensity ^21 22 23^. Progressive tissue destruction in chronic lesions was measured as an increase of MD or decrease of T1 intensity between baseline and follow-up timepoints^19 24^ using the baseline lesion mask, which was adjusted to correct for brain atrophy-related displacement of lesions at follow-up, as described previously ^3^.

**Suppl. eFig. 1.**
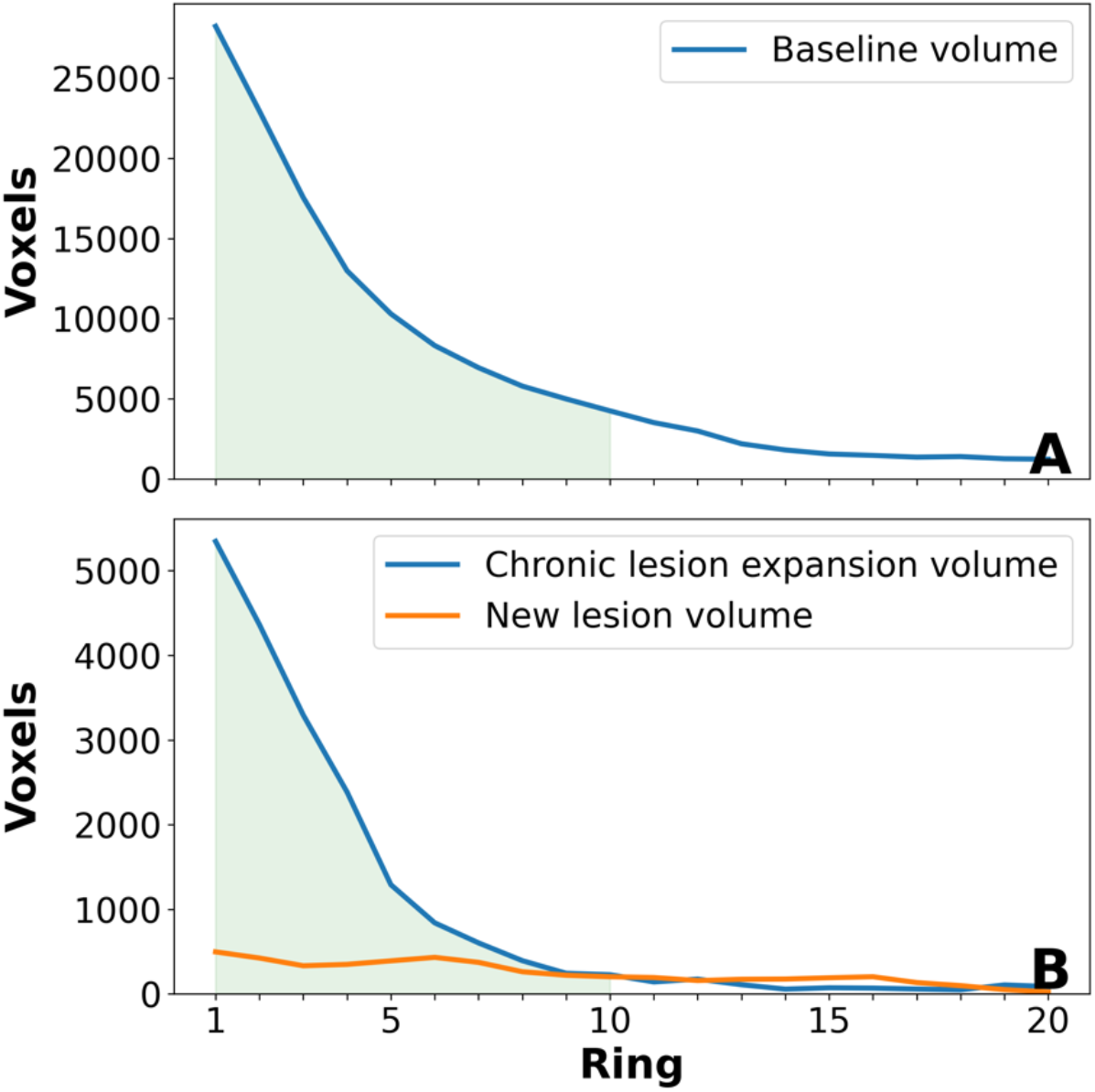
Ring-wise distribution of baseline lesion volume (A) and lesion change during follow-up (B). Green area marks lesion volume within the 10 periventricular rings analysed in this study.

**Suppl. eFig. 2.**
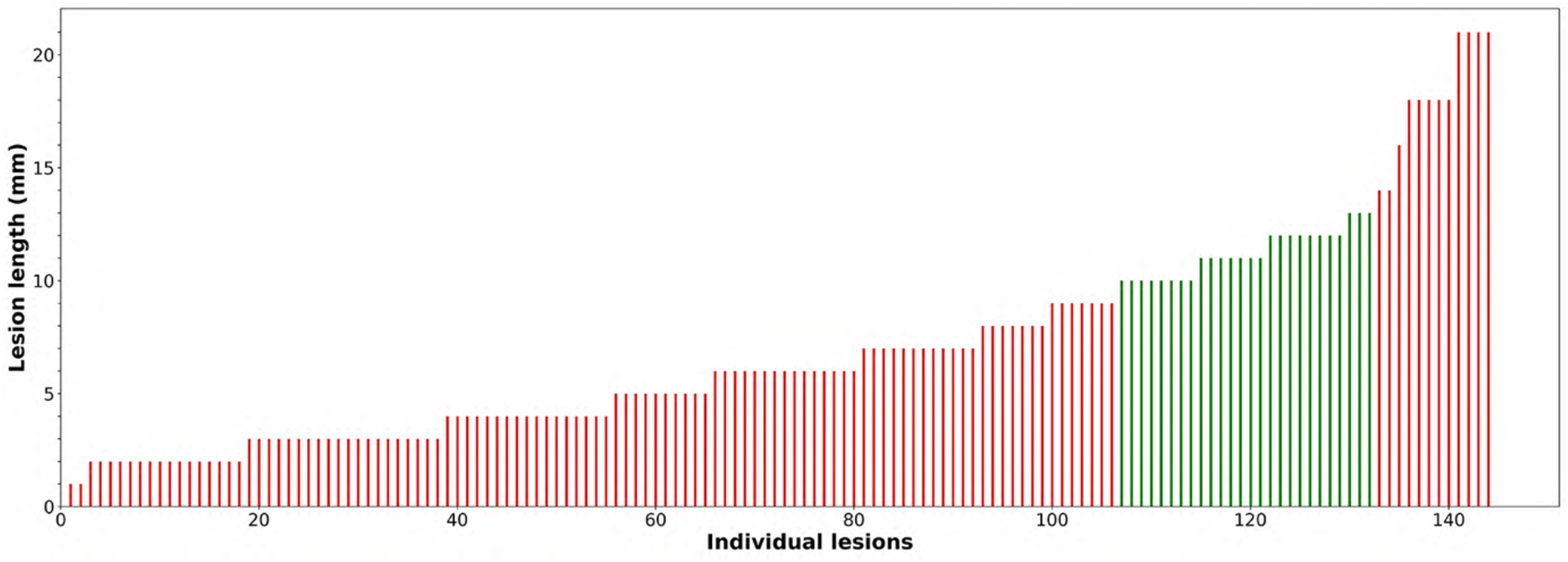
Distribution of lesion “length” for lesions adjacent to the ventricles

